# SARS-CoV-2 seroprevalence in children, parents and school personnel from June 2020 to April 2021: cohort study of 55 schools in Switzerland

**DOI:** 10.1101/2022.01.25.22269827

**Authors:** Agne Ulyte, Sarah R. Haile, Jacob Blankenberger, Thomas Radtke, Milo A. Puhan, Susi Kriemler

## Abstract

We measured SARS-CoV-2 seroprevalence in cohorts of children, parents and school personnel in 55 randomly selected schools in the canton of Zurich, Switzerland. In June-September 2020, seroprevalence was low (4.4% to 5.8%) in all cohorts. In March-April 2021, seroprevalence in children and parents (18.1% and 20.9%) was slightly higher than in school personnel (16.9%). Children’s seroprevalence was slightly higher in classes with infected main teachers and families with one infected parent, and substantially higher in families with two infected parents.

## Seroprevalence in children, school personnel and parents

*Ciao Corona* is a cohort study of 55 randomly selected schools from all districts of the canton of Zurich, Switzerland; described in detail earlier (7,8). Within participating schools, children from randomly selected classes, all school personnel and parents of participating children were invited.

Serology was measured in June-September 2020 (children: June-July; adults: August-September) and March-April 2021, with SenASTrIS test (Sensitive Anti-SARS-CoV-2 Spike Trimer Immunoglobulin Serological, sensitivity 98.3%, specificity 98.4%, as determined in population based samples) (9,10). We excluded vaccinated adults (n=123) and those eligible for vaccination but with unclear vaccination status (n=215) from the main analysis.

Table 1 presents age, sex and serological status of participants.

**Table 1.**
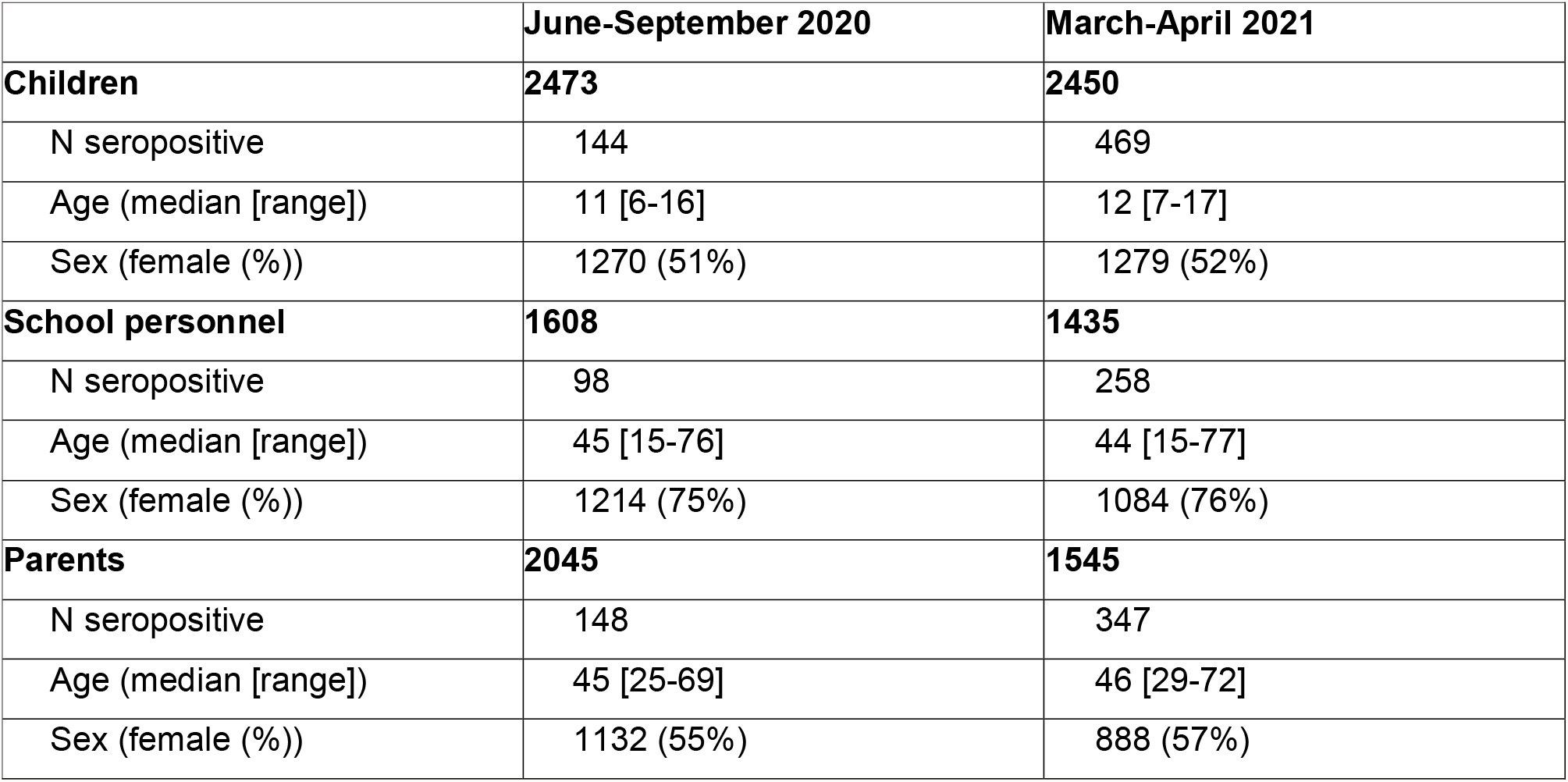
Characteristics of the children, school personnel and parent participant cohorts in June-September 2020 and March-April 2021

We adjusted seroprevalence estimates for test accuracy using parametric bootstrapping sampling (11,12). In June-September 2020, the adjusted seroprevalence was 4.4% (95% confidence interval 2.1-6.2%) among children, 4.6% (2.3-6.7%) among school personnel, and 5.8% (3.5-7.8%) among parents.

In March-April 2021, seroprevalence was 18.1% (15.7-20.7%) among children, 16.9% (14.1-19.8%) among school personnel, and 20.9% (18.0-23.9%) among parents. Seroprevalence was slightly lower (15.0% in school personnel and 19.3% in parents) when incorporating adjusted results from vaccinated and potentially vaccinated adults (Appendix 1).

## Variation in seroprevalence across districts and school communities

We further explored the variability of seroprevalence within and across cantonal districts and school communities in March-April 2021. We defined school community as a single school or a pair of a primary and a secondary school serving the same area. We compared seroprevalence with the per capita cumulative incidence of confirmed SARS-CoV-2 infections in districts and the postal codes constituting tested school communities, weighted by the number of tested children residing in them.

The relationship between the seroprevalence of children, school personnel, and parents’ cohorts within districts and school communities was not consistent (Figure 1), with large variation in cohort estimates also across districts and school communities.

**Figure 1.**
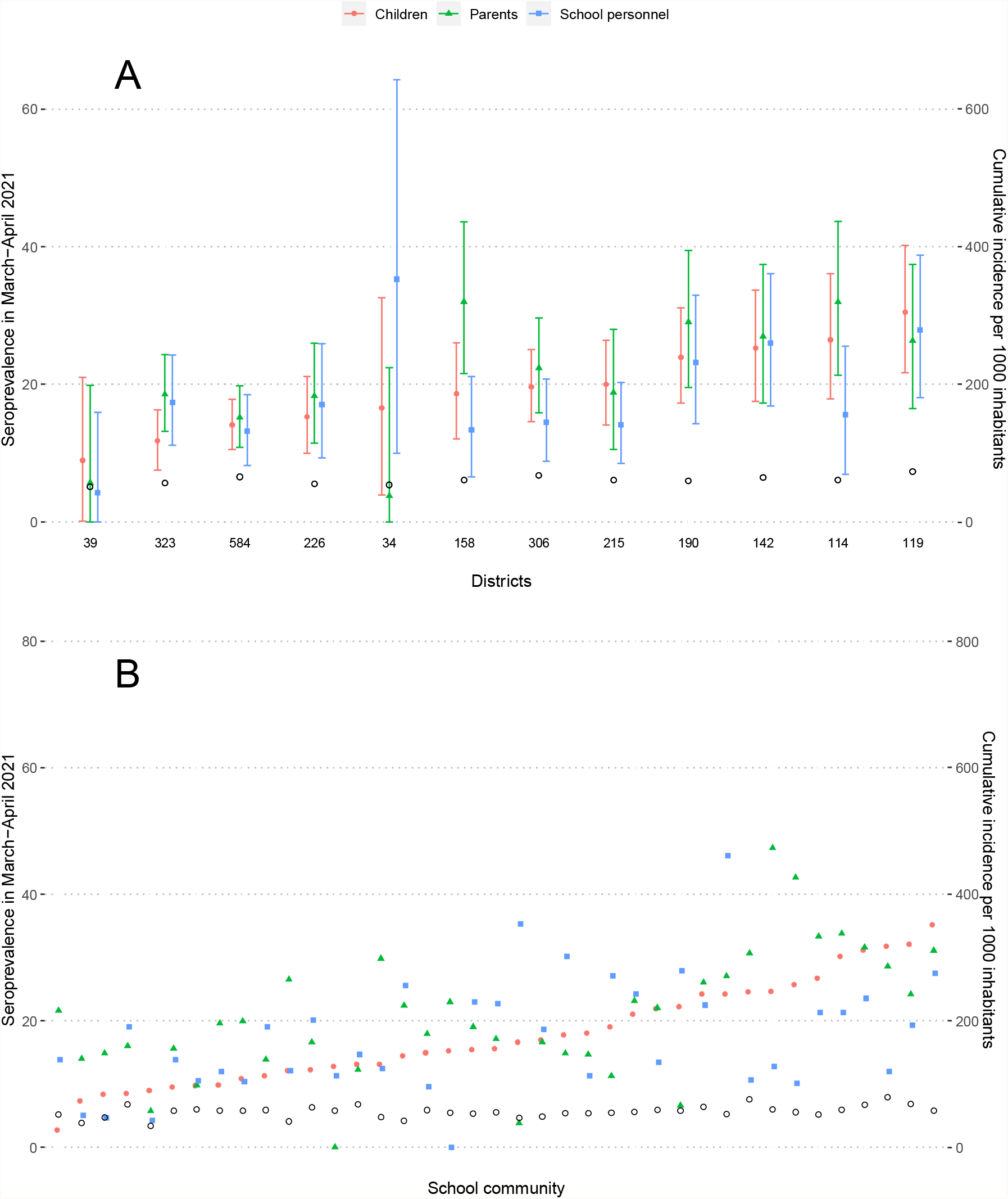
Seroprevalence in children, parents and school personnel and cumulative incidence of the population in the canton of Zurich, Switzerland, in March-April 2021 A – within the cantonal districts, B – within school communities Empty black circles represent cumulative incidence of laboratory-confirmed SARS-CoV-2 infections among inhabitants of the district by the middle of the testing period. At the bottom of Figure 1A are numbers of children tested within the district. Districts and school communities are ordered by increasing seroprevalence in children.

Median difference between parent and children seroprevalence in districts was 2.2% (interquartile range (IQR) -1.7% to 5.2%), and between school personnel and children -1.8% (IQR -5.2% to 1.0%).

Median difference between parents’ and children’s seroprevalence in school communities was 2.9% (IQR -1.8% to 7.1%), and between school personnel and children -0.5% (IQR -7.2% to 7.3%). Distributions were similar when analyzing children (N=577) and parents (N=1016) only from families in which both parents were tested (Appendix 2).

Variation in seroprevalence of SARS-CoV-2 was more pronounced than variation in cumulative incidence. The median ratio of children’s seroprevalence to per capita confirmed cases in district inhabitants was 3.1 (range 1.8 to 4.3, IQR 2.6 to 3.9).

## Seroprevalence by sex, school level and school personnel job

We observed no sex differences in the seroprevalence within cohorts (Table 2). Children and parents with children in upper school level had lower seroprevalence. There was no substantial difference in seroprevalence of children with tested and non-tested parents (18.5% vs 17.9%). Among school personnel, teachers and canteen personnel were seropositive most frequently (17.8%, N=1133, and 17.2%, N=88), followed by cleaning personnel (15.3%, N=67) and school administration (11.5%, N=126).

**Table 2.** Seroprevalence with 95% confidence interval in children, parents and school personnel based on sex and school level in March-April 2021

|  | Children |  | Parents<sup>a</sup> |  | School personnel<sup>b</sup> |  |
| --- | --- | --- | --- | --- | --- | --- |
|  | N | Seroprevalence | N | Seroprevalence | N | Seroprevalence |
| **Sex** |  |  |  |  |  |  |
| Female | 1279 | 18.2% (15.3-21.3) | 888 | 21.5% (18.1-25.1) | 1084 | 16.7% (13.9-20.1) |
| Male | 1165 | 18.1% (15.1-21.2) | 645 | 20.2% (16.4-24.1) | 347 | 17.1% (12.5-22.0) |
| **School level** |  |  |  |  |  |  |
| Lower | 768 | 18.1% (14.7-21.7) | 669 | 22.6% (18.8-26.7) | 216 | 21.3% (15.3-27.9) |
| Middle | 845 | 20.3% (16.9-23.9) | 599 | 23.4% (19.4-27.7) | 181 | 17.2% (11.2-24.0) |
| Upper | 837 | 16.0% (12.7-19.3) | 392 | 17.8% (13.4-22.6) | 227 | 18.4% (12.8-24.5) |

a - Parents with tested children in several school levels are included in all relevant groups.
b - Only class teachers are included in the estimates stratified by school level. Teachers in several school levels are included in all relevant groups. Seroprevalence in non-teaching school personnel was 15.6% (12.4-18.9) [N=864].

## Serological results within classes and families

In March-April 2021, children from 275 classes (median 5 classes per school) were tested. In 134 classes, at least 4 children, at least 25% of the children and at least one main teacher (teaching the tested class ≥14 hours per week for ≥ 6 months in 2020/2021) were tested. Among these, classes with seropositive main teachers had higher proportion of seropositive children (Figure 2A). For 1126 (46%) of children, at least one parent was tested; two parents were tested for 577 (24%) children. Children with two seropositive parents were the most likely to be seropositive, followed by children with a single seropositive parent (Figure 2B).

**Figure 2.**
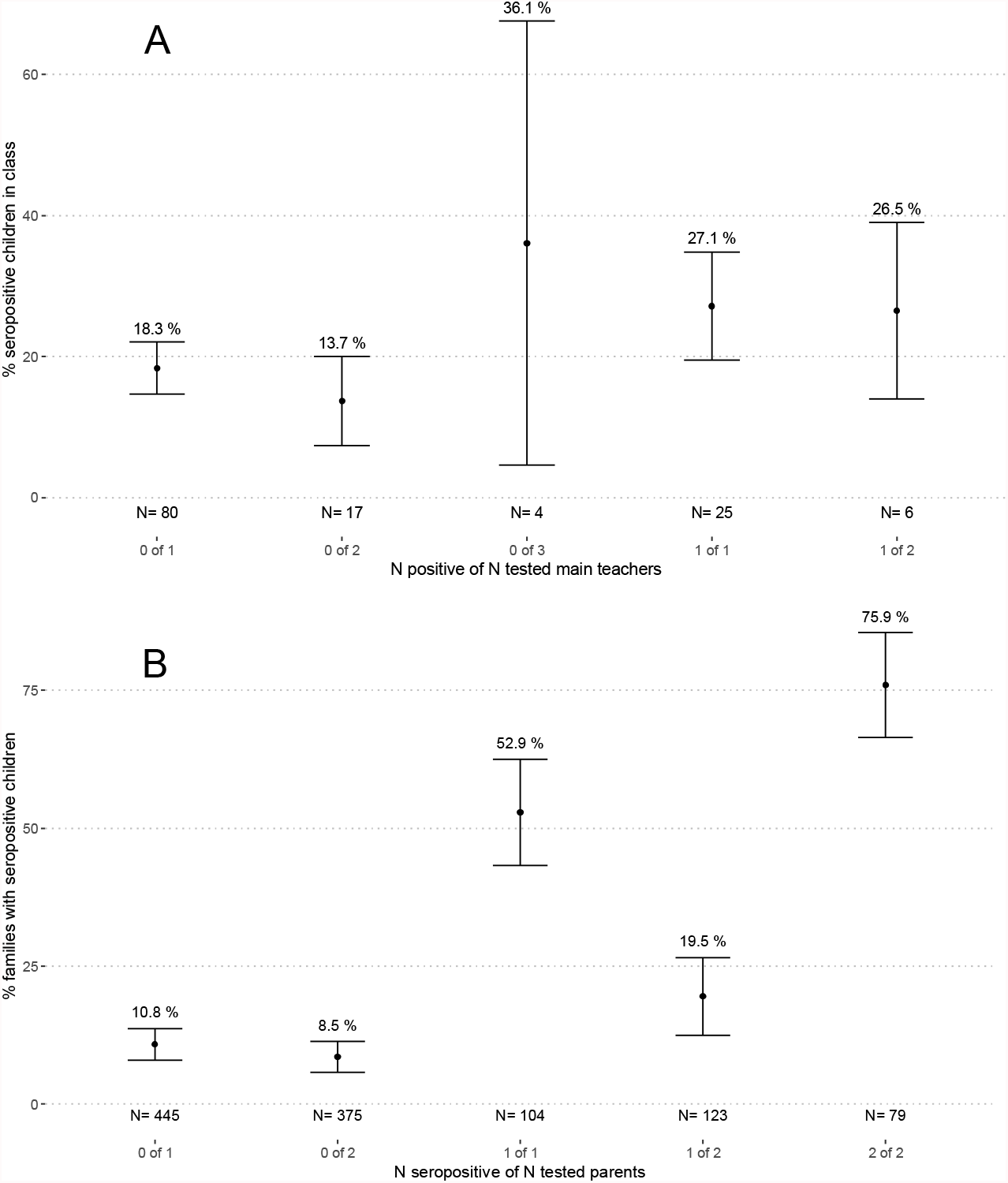
Proportion of seropositive children in classes and families depending on the number of tested and seropositive main teachers and parents in March-April 2021 A – classes with at least one main teacher and 4 children tested. Numbers (N) below graphs correspond to the numbers of classes in the category. B – families with at least one child and one parent tested. Numbers (N) below graphs correspond to the numbers of families in the category. Note: the name of the category, listed below the number, tells how many teachers (parents) were tested in a class (family) from the total tested within that class (family). E.g., “0 of 1” signifies classes where a single main teacher was tested, with test result seronegative. We do not show two classes with 3 main teachers tested, as confidence intervals could not be estimated for single data points.

## Discussion and conclusions

The seroprevalence of SARS-CoV-2 in June-September 2020 was low in children, parents and school personnel. Seroprevalence increased significantly by March-April 2021, and was slightly higher in parents and children than in school personnel. The pattern varied considerably within and between districts and school communities. We observed two-fold variation in the ratio of seropositive children to the cumulative incidence of confirmed SARS-CoV-2 infections in districts. This highlights that relying on the PCR-confirmed SARS-CoV-2 infections to estimate the spread of SARS-CoV-2 has drawbacks – and further stresses the importance of representative population-based seroprevalence studies.

Children and parents had lower seroprevalence in families with children in upper school levels; this trend was not observed in school personnel. All school personnel and children in upper school level were consistently wearing masks from November 2020. In contrast, masks were not implemented in lower school level and only for a few months in middle school level children (13). On the class level, classes with a main teacher testing seropositive had on average higher proportion of seropositive children compared to those with seronegative teacher. However, the difference was not large (27% vs 19%). In contrast, differences in proportions of seropositive children in families with 0, 1 and 2 seropositive parents were much larger (9% vs 20% vs 76%).

This study presents unique combined results of children, their parents and teachers from the same school communities and districts, and adds to our understanding of the mostly undetected infection chains in schools and families. However, it has several limitations. First, despite random sampling of schools and classes, children participating within classes (approximately 50%) might be somewhat (self) selected. Parents were more likely to attend testing in March-April 2021 if their child reported symptoms in 2020-2021 (at least one parent of 52% asymptomatic and 67% symptomatic children participated). However, participation was not different in parents of seropositive and seronegative children. Second, although serological tests have the advantage of capturing also the non-diagnosed SARS-CoV-2 infections, temporal and directional associations cannot be deduced. Positive serological test result could reflect an infection of up to a year ago, although antibodies are subject to some waning (14). Finally, the study was conducted when the delta or omicron variants of SARS-CoV-2 were not yet prevalent in Switzerland. Due to higher infectiousness, the associations of seroprevalence within families and classes might be different beyond the timeframe of this study.

In conclusion, we observed similar seroprevalence in children and parents, and somewhat lower in school personnel in March-April 2021 in Zurich, Switzerland, and striking variation between districts and school communities. Children’s seroprevalence was higher in classes with infected main teachers and from families with infected parent(s). The class and school community trends in seroprevalence could also be driven by community transmission outside and around rather than inside the school.

## Data Availability

Data is still being collected for the cohort study Ciao Corona. Deidentified participant data might be available on reasonable request by email to the corresponding author at later stages of the study.

## Funding statement

This study is part of *Corona Immunitas* research network, coordinated by the Swiss School of Public Health (SSPH+), and funded by fundraising of SSPH+ that includes funds of the Swiss Federal Office of Public Health and private funders (ethical guidelines for funding stated by SSPH+ will be respected), by funds of the Cantons of Switzerland (Vaud, Zurich and Basel) and by institutional funds of the Universities. Additional funding, specific to this study is available from the University of Zurich Foundation.

The spread of severe acute respiratory syndrome coronavirus 2 (SARS-CoV-2) in schools has been variable across regions and at different time points (1–4). Few studies have directly examined incidence or seroprevalence of SARS-CoV-2 infections in children, parents, and teachers from the same school communities (5,6).

Despite high incidence of SARS-CoV-2 in Switzerland, children attended school in person continuously in 2020-2021 except for 6 weeks in March-May 2020, which is quite exceptional internationally. The aim of this study is to describe SARS-CoV-2 seroprevalence within cantonal districts and school communities in children, parents and school personnel in June-September 2020 and March-April 2021. The study was approved by the Ethics Committee of the Canton of Zurich, Switzerland (2020-01336). Participants (or their parents) provided written informed consent.

## Acknowledgements

We would like to thank all participating children, parents and school personnel. We would like to thank Priska Ammann for organizing the March-April 2021 testing round of the study.

## Author contributions

Agne Ulyte: study design, data collection, analysis, interpretation, manuscript writing

Sarah R. Haile: study design, data analysis, manuscript revision

Jacob Blankenberger: study design, data collection, manuscript revision

Thomas Radtke: study design, data collection, interpretation, manuscript revision

Milo A. Puhan: study design, data collection, interpretation, manuscript revision

Susi Kriemler: study design, data collection, interpretation, manuscript revision

