## Supplementary materials for "SARS-CoV-2 seroprevalence in children, parents and school personnel from June 2020 to April 2021: cohort study of 55 schools in Switzerland"

**Appendix 1** Study results including the vaccinated parents and school personnel

Appendix 1 incorporates in the analysis of the participant cohorts also the results of vaccinated and potentially vaccinated parents and school personnel.

By April 2021 in Switzerland, persons over 50 years as well as some high-risk populations were eligible for SARS-CoV-2 vaccination. At the time of testing in 2021, approximately 8% of parents and 3% of school personnel were vaccinated against SARS-CoV-2. In interpreting their serological results, we considered self-reported vaccination status and detection of IgG against SARS-CoV-2 antigen N (N-IgG).

Participating adults negative for S-IgG and IgA were considered seronegative. Adults positive for S-IgG or IgA and positive for N-IgG were considered seropositive due to infection regardless of vaccination status. Vaccinated adults positive for S-IgG or IgA and negative for N-IgG were considered seropositive due to vaccination only, and further analysed as seronegative. We removed participants with both uncertain vaccination information and negative N-IgG from analysis (3 school personnel and 7 parents). In this Appendix, we further refer to as "seropositive" only persons with positive serology due to SARS-CoV-2 infection.

Table S1 describes the characteristics of the participants.

**Table S1** Characteristics of the children, school personnel and parent participant cohorts

| | **March-April 2021** |
|---|---|
| **Children** | **2450** |
| N seropositive | 496 |
| Age (median [range]) | 12 [7-17] |
| Sex (female (%)) | 1279 (52%) |
| **School personnel** | **1604** |
| N seropositive | 259 |
| Age (median [range]) | 45 [15-77] |
| Sex (female (%)) | 1215 (76%) |
| **Parents** | **1714** |
| N seropositive | 347 |
| Age (median [range]) | 46 [29-71] |
| Sex (female (%)) | 992 (58%) |

In March-April 2021, the adjusted seroprevalence was 18.1% (15.7-20.7%) among children, 15.0% (12.4-17.7%) among school personnel, and 19.3% (16.6-22.1%) among parents. The adjusted seroprevalence in the cantonal districts is shown in Figure S1A, along with the cumulative incidence of confirmed SARS-CoV-2 infections. The relationship between the seroprevalence of the three cohorts within districts and school communities was not consistent.

**Figure S1** Seroprevalence in children, parents and school personnel and cumulative incidence of the population in the canton of Zurich, Switzerland, in March-April 2021

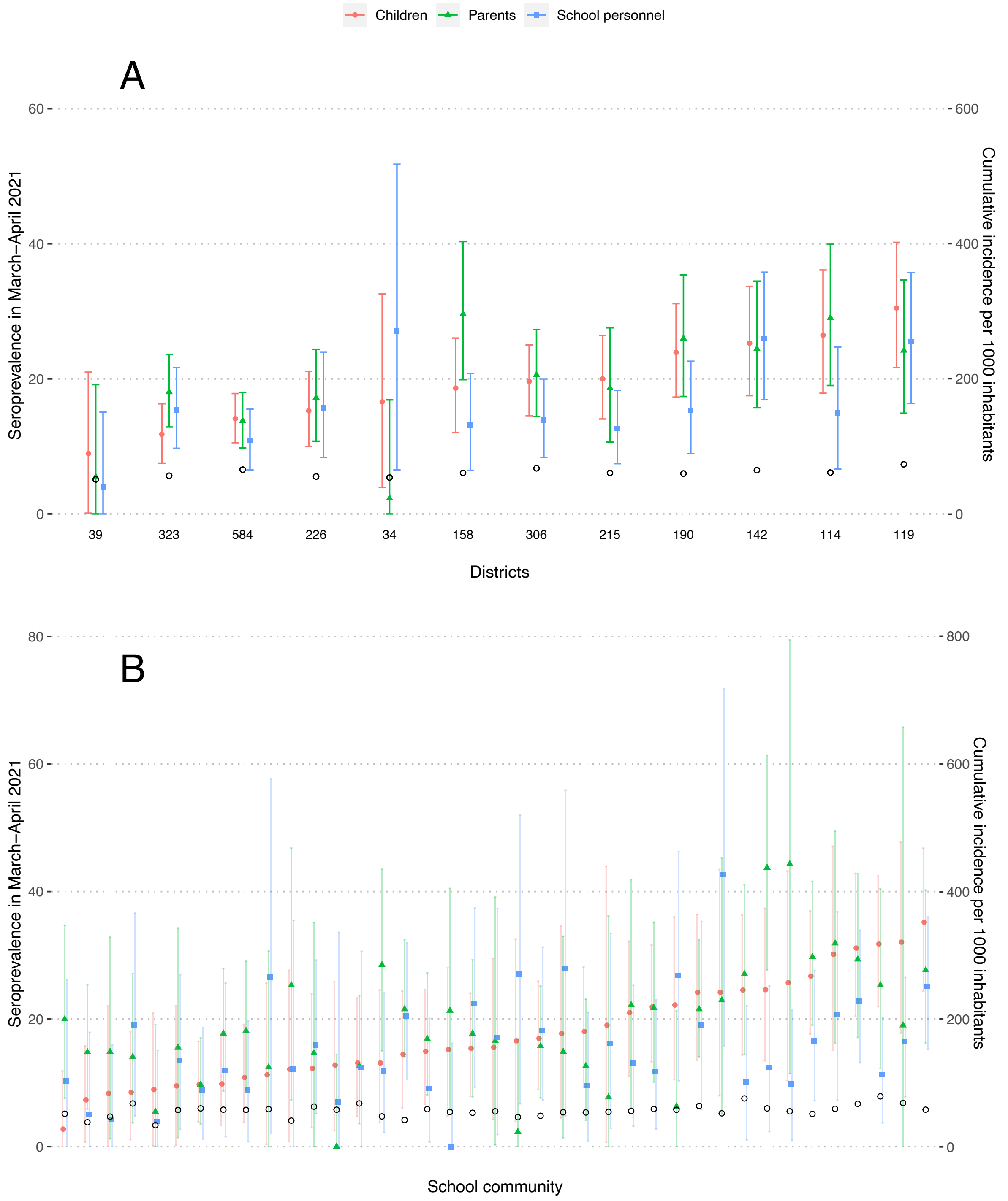


A – within the cantonal districts, B – within school communities

Empty black circles represent cumulative incidence of laboratory-confirmed SARS-CoV-2 infections among inhabitants of a district. At the bottom of Figure 1A are numbers of children tested within the district. Districts and school communities are ranked by increasing seroprevalence in children.

Median difference between parent and children seroprevalence in districts was 0% (interquartile range (IQR) -1.9% to 1.7%), and between school personnel and children -5.0% (IQR -6.2% to 0.5%).

Median difference between parent and children seroprevalence in school communities was 1.1% (IQR -2.7% to 6.3%), and between school personnel and children -2.3% (IQR -8.9% to 3.8%).

No sex differences were observed in the seroprevalence within cohorts (Table S2). There were no differences in seroprevalence of children with tested and non-tested parents (18.5% vs 17.8%). Teachers and canteen personnel were seropositive most frequently (16.1%, N=1247, and 15.4%, N=97), followed by cleaning personnel (11.9%, N=84) and school administration (9.2%, N=152). Children and parents with children in upper school level had lower seroprevalence.

**Table S2** Seroprevalence with 95% confidence interval [number of participants] in children, parents and school personnel based on sex and school level in March-April 2021

| | Children | | Parents[a] | | School personnel[b] | |
|---|---|---|---|---|---|---|
| | N | Seroprevalence | N | Seroprevalence | N | Seroprevalence |
| **Sex** | | | | | | |
| Female | 1279 | 18.2% (15.3-21.3) | 992 | 19.8% (16.6-23.2) | 1215 | 14.9% (12.1-17.8) |
| Male | 1165 | 18.1% (15.1-21.2) | 708 | 18.7% (15.1-22.4) | 385 | 15.5% (11.2-20.1) |
| **School level** | | | | | | |
| Lower | 768 | 18.1% (14.7-21.7) | 711 | 21.0% (17.4-24.9) | 240 | 19.0% (13.5-25.1) |
| Middle | 845 | 20.3% (16.9-23.9) | 660 | 21.5% (17.7-25.5) | 193 | 16.0% (10.2-22.4) |
| Upper | 837 | 16.0% (12.7-19.3) | 448 | 15.9% (11.8-20.2) | 245 | 17.3% (12.0-23.1) |

a - Parents with tested children in several school levels are included in all relevant groups.

b - Only class teachers are included in the estimates stratified by school level. Teachers in several school levels are included in all relevant groups. Seroprevalence in non-teaching school personnel was 13.5% (10.5-16.5) [N=983].

In 141 classes, at least 4 children and at least 25% of the class children as well as at least one main teacher were tested. Classes with seropositive main teachers had higher proportion of seropositive children (Figure S2A). At least one parent was tested for 1202 (49%) of the tested children, and two parents for 674 (28%) of the tested children. Children with two seropositive parents were the most likely to be seropositive, followed by children with a single seropositive parent (Figure S2B).

**Figure S2** Proportion of seropositive children in classes and families depending on the number of tested and seropositive main teachers and parents in March-April 2021

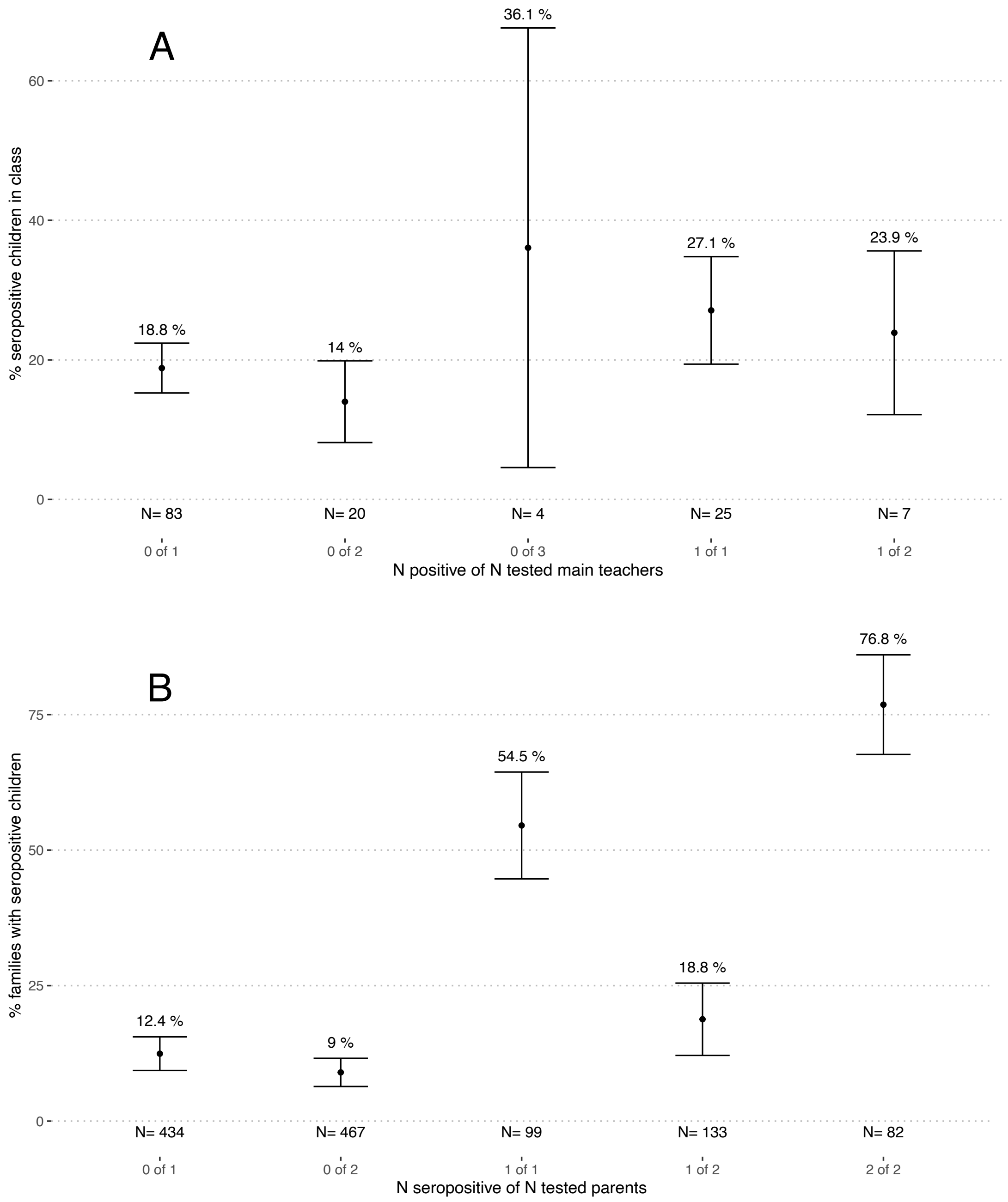


A – classes with at least one main teacher and 4 children tested. Numbers (N) below graphs correspond to the numbers of classes in the category.

B – families with at least one child and one parent tested. Numbers (N) below graphs correspond to the numbers of families in the category.

Note: the name of the category, listed below the number, tells how many teachers (parents) were tested in a class (family) from the total tested within that class (family). E.g., “0 of 1” signifies classes where a single main teacher was tested, with test result seronegative. We do not show two classes with 3 main teachers tested, as confidence intervals could not be estimated for single data points.

### Appendix 2

**Figure S3** Seroprevalence in children, parents and school personnel and cumulative incidence of the population in the canton of Zurich, Switzerland, in March-April 2021. Only children and parents from families in which both parents were tested are included.

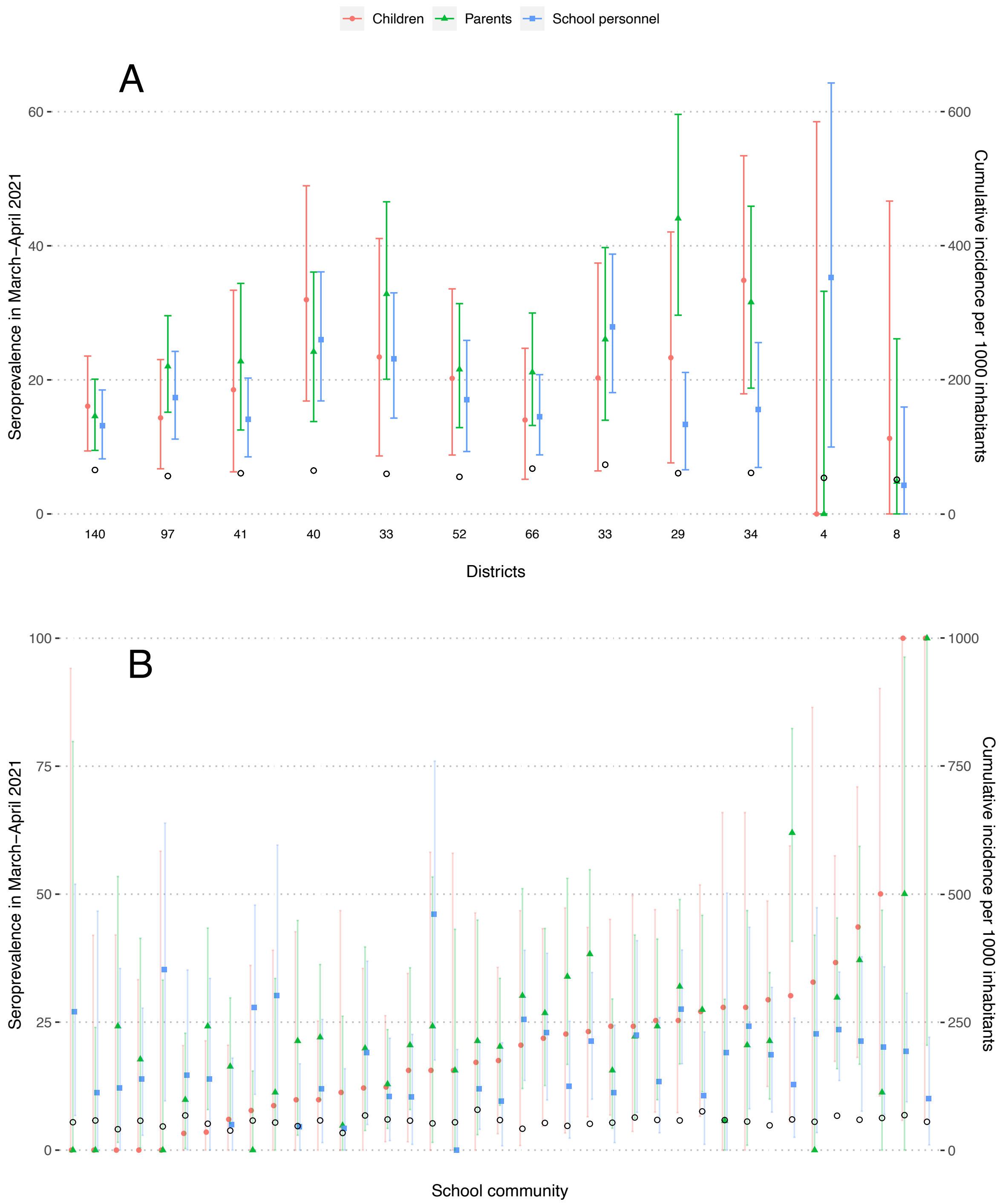


A – within the cantonal districts, B – within school communities

Empty black circles represent cumulative incidence of laboratory-confirmed SARS-CoV-2 infections among inhabitants of the district. At the bottom of Figure 1A are numbers of children tested within the district. Districts and school communities are ordered by increasing seroprevalence in children.

Median difference between parent and children seroprevalence in districts was 2.8% (interquartile range (IQR) -1.9% to 7.3%), and between school personnel and children -3.0% (IQR -6.2% to 1.1%).

Median difference between parent and children seroprevalence in school communities was 0.6% (interquartile range (IQR) -6.4% to 9.1%), and between school personnel and children -3.7% (IQR -11.3% to 8.6%).
